# Correlates of suicidal behaviors and genetic risk among United States veterans with schizophrenia or bipolar I disorder

**DOI:** 10.1101/2023.03.06.23286866

**Authors:** Tim B. Bigdeli, Peter B. Barr, Nallakkandi Rajeevan, David P. Graham, Yuli Li, Jacquelyn L. Meyers, Bryan R. Gorman, Roseann E. Peterson, Frederick Sayward, Krishnan Radhakrishnan, Sundar Natarajan, David A. Nielsen, Anna V. Wilkinson, Anil K. Malhotra, Hongyu Zhao, Mary Brophy, Yunling Shi, Timothy J. O’Leary, Theresa Gleason, Ronald Przygodzki, Saiju Pyarajan, Sumitra Muralidhar, J. Michael Gaziano, Million Veteran Program (MVP), Cooperative Studies Program (CSP) #572, Grant D. Huang, John Concato, Larry J. Siever, Lynn E. DeLisi, Nathan A. Kimbrel, Jean C. Beckham, Alan C. Swann, Thomas R. Kosten, Ayman H. Fanous, Mihaela Aslan, Philip D. Harvey

## Abstract

**Objective:** Persons diagnosed with schizophrenia (SCZ) or bipolar I disorder (BPI) are at high risk for self-injurious behavior, suicidal ideation, and suicidal behaviors (SB). Characterizing associations between diagnosed mental and physical health problems, prior pharmacological treatments, and aggregate genetic factors has potential to inform risk stratification and mitigation strategies.

**Methods:** In this study of 3,942 SCZ and 5,414 BPI patients receiving VA care, self-reported SB and ideation were assessed using the Columbia Suicide Severity Rating Scale (C-SSRS). These cross-sectional data were integrated with electronic health records (EHR), and compared by lifetime diagnoses, treatment histories, follow-up screenings, and mortality data. Polygenic scores (PGS) for traits related to psychiatric disorders, substance use, and cognition were constructed using available genomic data, and exploratory genome-wide association studies were performed to identify and prioritize specific loci.

**Results:** Only 20% of veterans who self-reported SB had a corroborating ICD-9/10 code in their EHR; and among those who denied prior behaviors, more than 20% reported new-onset SB at follow-up. SB were associated with a range of psychiatric and non-psychiatric diagnoses, and with treatment with specific classes of psychotropic medications (e.g., antidepressants, antipsychotics, etc.). PGS for externalizing behaviors, smoking, suicide attempt, and major depressive disorder were also associated with attempt and ideation.

**Conclusions:** Among individuals with a diagnosed mental illness, a GWAS for SB did not yield any significant loci. Self-reported SB were strongly associated with clinical variables across several EHR domains. Overall, clinical and polygenic analyses point to sequelae of substance-use related behaviors and other psychiatric comorbidities as strong correlates of prior and subsequent SB. Nonetheless, past SB was frequently not documented in clinical settings, underscoring the value of regular screening based on direct, in-person assessments, especially among high-risk individuals.

## INTRODUCTION

Suicide is the leading cause of injury-related mortality in the United States, surpassing motor vehicle accidents, homicide, and opiate overdoses, and is the tenth overall cause of death(1). Deaths by suicide have increased 33% over the past two decades and are increasing more rapidly in veterans than in non-veterans(2). Additionally, this increase in deaths by suicide has been attributed as one of the causes of recent decreases in life expectancy, alongside other “deaths of despair” (e.g., drug-related and alcohol related deaths)(3). Given that immediate suicidal behaviors (SB) are often unpredictable, with death commonly occurring on the first attempt, and because no medicines have been shown to widely reduce deaths by suicide (outside of clozapine in patients with schizophrenia), there is a critical need for new strategies to identify persons at high risk before their first attempt.

Risk factors for suicide include a variety of social, clinical, and biological correlates. Demographic factors, including age, gender, race, ethnicity, marital status, and socioeconomic status, are associated with higher risk of suicide(4). Attempted suicides are much more common among persons with diagnosed mental illnesses such as major depressive disorder (MDD), substance-use disorders (SUD), and posttraumatic stress disorder (PTSD), and even more markedly so, schizophrenia (SCZ) and bipolar I and II disorder (BPI). In addition to phenotypic correlations, a growing body of evidence supports a partial, shared genetic basis of SB with a range of psychiatric traits and disorders(5–9).

United States (US) veterans who receive services from the Veterans Health Administration (VHA) commonly have histories of trauma exposure, both combat- and noncombat-related, and posttraumatic stress disorder (PTSD), smoking, cardiovascular disease, certain cancers, and suicide. For more than a decade, the VHA has facilitated research aimed at improving health outcomes among these Veterans, including the Million Veteran Program(10) and a companion study of 9,300 veterans with confirmed serious mental illnesses, Cooperative Studies Program (CSP) #572, *The Genetics of Functional Disability in Schizophrenia (SCZ) and Bipolar Illness (BPI)(11)*. In addition to expansive clinical data available from the VHA electronic health record (EHR), a unique strength of CSP #572 is its detailed in-person assessments of each Veteran, which included the Columbia Suicide Severity Rating Scale (C-SSRS)(12). This can be used to clarify diagnoses that can be uncertain, incomplete, or even biased, when based on EHR alone(13)

Our previous studies from CSP #572(14, 15) highlighted veterans with serious mental illnesses (SMI) as a special population at particularly high risk of suicide, 46% and 55% of SCZ and BPI patients, respectively, reported past SB, with only 30% and 18% denying lifetime ideation or behavior. Factors associated with increased risk for SB included being female, younger age, a BPI diagnosis, higher cognitive performance, lower income and education attainment, having been married, and a greater number of comorbidities. Drug use, alcohol use, and being a smoker were associated with higher short-term all-cause mortality in the six years following completion of study enrollment.

To better understand how suicide and suicide-related risk factors are characterized in EHRs for persons with severe mental illnesses, we combined information from detailed in-person assessments for CSP #572 with ICD-9/10 billing codes, prescription data, and common laboratory values from EHR. First, we evaluated whether participants’ C-SSRS data from a clinical interview were corroborated by relevant ICD-9/10 billing codes and employed both phenome-wide association studies (PheWAS) and clinical laboratory-wide association studies (LabWAS) to explore phenotypic correlations with SB more broadly. Next, using available genomic data, we evaluated associations between SB and polygenic scores (PGS) for known correlates of suicide and performed both genome-wide association studies (GWAS) and transcriptome-wide association studies (TWAS) to identify biologically plausible risk loci. Finally, we compared baseline data for CSP #572 alongside information from follow-up C-SSRS screenings in routine health care visits and National Death Index (NDI) records.

## METHODS

### Participants

CSP 572 was approved by the VA Central IRB, and all participants provided written informed consent. No participants requiring the permission of a guardian to participate were enrolled.

Participants were recruited under the oversight of a site PI and research assistants, with the aid of clinicians providing care to patients with SCZ and BPI. Advertisements about the study were also in public locations at 27 participating VA hospitals. Veterans who participated also informed peers and encouraged them to participate. Sites were selected for CSP 572 based on criteria including previous successful participation in VA research on severe mental illness and the availability of large numbers of veterans with SMI diagnoses for recruitment (see Supplementary Acknowledgements).

All participants met lifetime DSM-IV criteria for SCZ (any type), or Bipolar I disorder, which requires a history of at least 1 mixed or full manic episode. Participants with major neurologic illnesses, medical problems that could interfere with central nervous system function, and those meeting criteria for schizoaffective disorder were excluded. Diagnosed substance use disorders were not an exclusion criterion, given a high co-occurrence in the study population. Participants were not enrolled if they appeared to be intoxicated at a study visit but could be reassessed later.

Information from medical charts, patients’ clinicians, or other informants was used, if needed, to confirm diagnoses, with all participants receiving the Structured Clinical Interview for the DSM-IV (SCID)(16). Consensus procedures, involving contact with a Clinical Study Chair (P.D.H.), were used to resolve diagnostic questions at local sites. Descriptive statistics for clinical and demographic factors are reported in Table S1.

### Direct Assessment of Lifetime Suicidality

Clinical assessment of suicidality used the C-SSRS(17) (version 1/14/09), which assesses suicidal ideation, suicide attempts, and their severity, allowing for the separation of suicide attempts from non-suicidal self-injurious acts. This stand-alone scale is straightforward regarding training and administration and is used in pharmacological treatment studies as a safety measure. We classified individuals as having exhibited lifetime SB if they endorsed any of the following: prior, aborted, or interrupted attempts, or preparatory acts. We classified individuals as having suicidal ideation (only) if they endorsed suicidal thoughts but denied any past SB. The first group of raters was trained in person by the developer of the scale and new raters received formal training and were certified.

In addition, we used the PTSD module from the MINI International Diagnostic interview (MINI-6)(18) to assess the presence of current PTSD (last 6 months) in all participants. Using the relevant section of the SCID, and only for SCZ patients, we assessed the lifetime history of any major depressive disorder (MDD) episode.

### Safety Plan

Assessments of both SB and ideation, as well as PTSD, in an already vulnerable population raised the concern of detecting potentially dangerous mental states in research participants. A detailed safety plan was developed, implemented, and monitored.

### Electronic Health Records (EHRs)

We extracted relevant ICD-9/10 billing codes indexing suicide attempts and suicidal ideation from the VA Corporate Data Warehouse (CDW) (Table S2). For comparisons of C-SSRS outcomes with **prior** diagnoses of suicidality, records were limited to those **predating** in-person assessments for CSP #572. In addition to these prior diagnoses of suicidality, 6,830 participants had at least one follow-up screening using the C-SSRS in their EHR. On average, these screenings occurred 8-12 years after their baseline assessment for CSP #572. Lastly, participants were linked to National Death Index (NDI) records that included cause of death.

We employed an analytical framework for phenome-wide association studies (PheWAS)(19) to summarize associations between SB (or ideation only) and “phecodes” representing groupings of related ICD-9/10 billing codes coming from the CDW(20). Recent analyses of phecodes within veteran populations show that two or more ICD-9/10 codes correspond strongly with structured clinical interviews(21). For PheWAS, we assumed a Bonferroni-adjusted significance threshold of P<10^−5^.

We also compiled a list of commonly prescribed antipsychotics, mood stabilizers, and antidepressants from the VA national formulary (Table S3) and extracted these from prescription records for CSP #572 participants, including date and dosage. All phecodes and medications were tested for association with C-SSRS data by logistic regression, covarying for age, sex, and specific diagnosis.

We created a proxy for comorbidity burden by tabulating the number of unique phecode terms for which each individual met our basic case/control criteria (i.e., 2 or more codes for cases, and zero for controls). Previous published analyses of co-morbidities and suicide outcomes in this sample (14, 15) had been restricted to calculation of the Charlson comorbidity index based on a health screening measure. We restricted this list to 600 “top codes”, excluding child terms or “leaf codes”, to limit over-representation of ontologically related terms. We tested the comorbidity index, within broad disease categories and overall, for association with SB, adjusting for the same covariates used in the primary PheWAS.

### Genotyping and Quality Control

Details of biospecimen collection, genomic data generation, and quality control have been detailed elsewhere(11, 13, 22). Briefly, specimens were processed together with those of enrollees from the MVP (www.research.va.gov/mvp) and genotyped on a customized Affymetrix Axiom Biobank array (Release 3). Duplicate samples and samples with excess missing genotype calls were excluded (>2.5%). Variants with >5% missingness or that deviated from their expected allele frequency (>10%) based on 1000 Genomes Project Phase 3 reference data (1000G)(23) were excluded. Imputation was performed using Minimac3(24).

Pairwise genetic relatedness was estimated using KING(25). Individuals demonstrating excess pairwise relatedness were excluded (KING coefficient > 0.177), and we removed one individual at random from each pair of first-degree relatives.

We applied the HARE algorithm(26) that incorporates both self-identified race/ethnicity and genetic data to assign individuals to broadly defined ancestry groups, namely admixed African (AA), European (EA), Hispanic, and East or South Asian. This general strategy has been shown to be robust to population stratification(13, 27, 28). Samples with excess heterozygosity outliers (>0.1) were identified within HARE groups and removed. Our genomic analyses are limited to veterans of admixed EA (N = 3,049) and AA (N = 2,285) with available genomic data.

Principal components analysis (PCA) was performed using flashpca2(29) within HARE groups based on a set of 27,071 LD-independent SNPs. Following visual inspection of PCA-based clustering, the first six ancestral principal components (PCs) were included as covariates in all genetic analyses (Figure S1).

### PGS and SNP-based heritability

We constructed PGS from published GWAS results of phenotypes related to SB (“training” datasets), testing individual-level scores for association with suicidality phenotypes in CSP #572 (“target” dataset). We included PGS for suicide attempt (30), psychiatric disorders (31–34), substance-related traits (9, 35), and cognitive functioning (36). Variants that met quality control requirements in both the original GWAS and the current analysis were clumped in the appropriate 1000G super population (*r*^2^>0.1; 500 kb window). For varying *P*-thresholds, scores were constructed by summing the number of tested alleles by its effect estimate. Differences were tested using logistic regression, covarying for age, sex, diagnosis, and six ancestry PCs.

We estimated the variance explained by the aggregate effects of genome-wide common single nucleotide polymorphisms (SNP), or SNP-*h*^2^, using genome-based restricted maximum likelihood (GREML), as implemented in the genome-wide complex trait analysis (GCTA) software (37, 38). We included age, sex, diagnosis, and six ancestry PCs as covariates and used the [--grm-cutoff .05] flag to restrict analyses to approximately unrelated individuals.

### Exploratory Genome-wide Association Studies

We conducted a variety of exploratory genomic analyses within our veteran sample. First, we performed a GWAS within CSP #572 participants (N = 5,334, 69.5% reporting lifetime attempt), testing for association between imputed genotype dosages and SB by logistic regression using PLINK(39, 40) including age, sex, diagnosis, and six ancestry PCs as covariates, and retaining variants with imputation quality ≥0.6 and MAF≥1%. After performing GWAS within ancestry groups, we combined results across using an inverse variance-weighted fixed-effects meta-analysis using METAL(41). We defined independent loci (P<10-6) by “clumping” of SNPs in LD (r2 ≥ 0.1) with the lead variant within a 3 megabase window (42). Clumped intervals within 250 kilobases were annealed into physically distinct loci. We used the Functional Mapping and Annotation (FUMA)(43) platform (http://fuma.ctglab.nl/) for bioinformatic annotation of independent loci, and LocusZoom(44) (http://locuszoom.org) for creating regional association plots.

In order to compare the results from this cohort with the broader GWAS literature on SB, we combined our results with single- and cross-disorder summary statistics from published meta-analyses of suicide attempts from the Psychiatric Genomics Consortium (PGC)(5) and International Suicide Genomics Consortium (ISGC) (30). We applied fixed effects meta-analyses to within-ancestry analyses, and Han and Eskin’s random effects model for multi-ancestry analyses; both are implemented in the METASOFT software package (45).

Lastly, we performed a TWAS to explore whether there was tissue-specific enrichment in gene expression for regions identified in the main GWAS. We utilized the S-PrediXcan software(46, 47) to predict gene expression differences from GWAS summary statistics, using prediction models for 13 CNS tissues and 3 endocrine tissues from the Genotype-Tissue Expression (GTEx v8) database (http://predictdb.hakyimlab.org/). False discovery rate (FDR) corrections of 5% were applied within and across tissues.

## RESULTS

### Prior suicidal behaviors are underreported in EHRs

Figure 1 and Tables S4 summarize numbers of participants who self-reported SB (N = 4,766), ideation (N = 2,445), or non-suicidal self-injury (N = 91), and the correspondence of C-SSRS assessments with ICD-9/10 billing codes for suicide attempts and suicidal ideation. Among participants self-reporting SB, fewer than 20% had a corroborating ICD-9/10 code for attempt or ideation in their EHR prior to their clinical interview. Similarly, fewer than 10% of those who endorsed ideation but denied behavior had any relevant codes. A similar pattern was observed for the ∼1% of participants who denied past SB and ideation but reported self-injurious behaviors.

**Figure 1.**
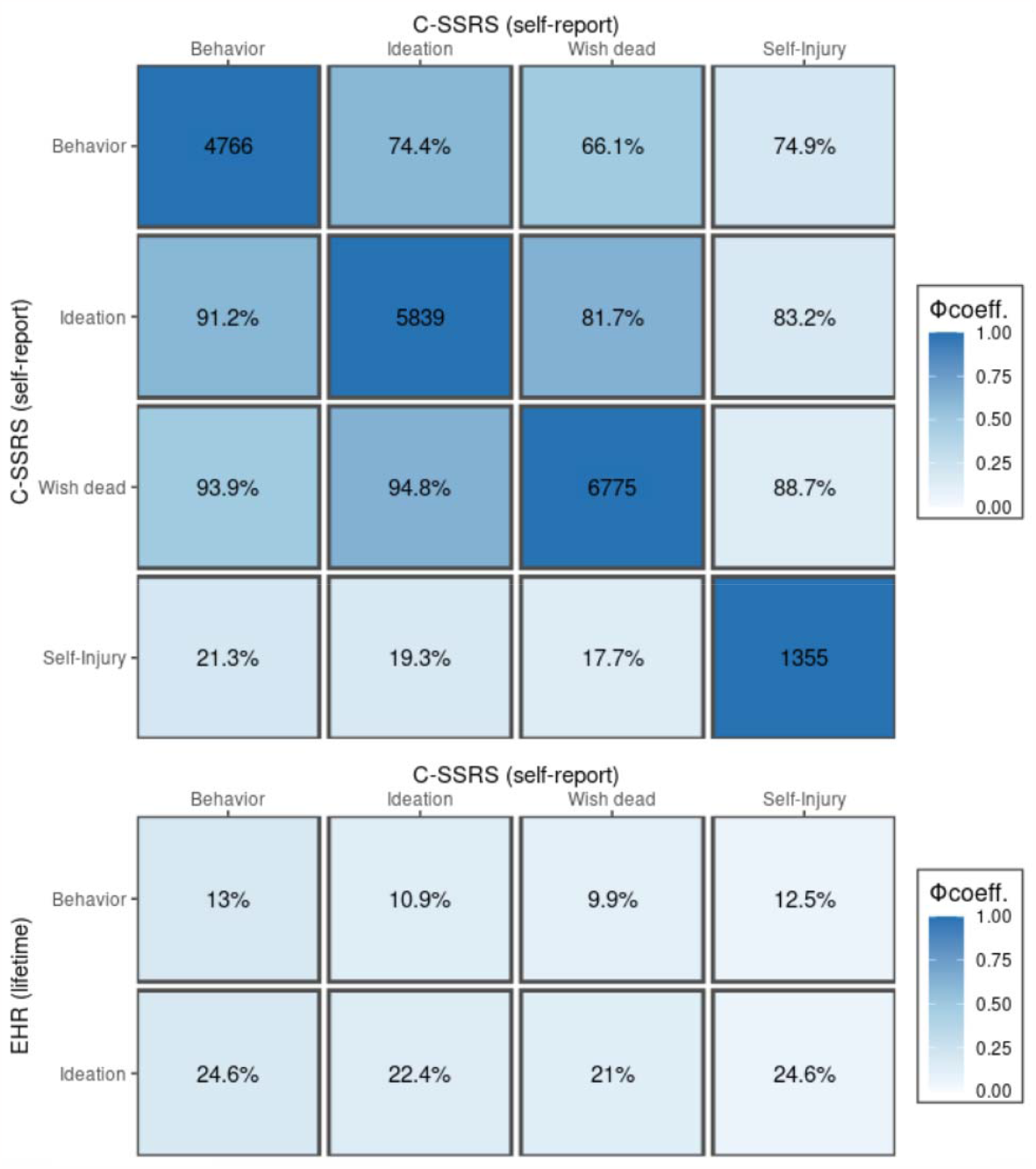
Phenotypic correlations for C-SSRS and EHR-based ratings of suicidality. (*top*) Numbers of participants endorsing each C-SSRS category are displayed on the diagonal; cross-tabulations for the proportion of participants meeting both criteria are displayed in the lower triangle (for columns) and upper triangle (for rows) as percentages; cells are shaded according to the Phi correlation coefficient (Φ) for dichotomous variables. (*lower*) as above but contrasting C-SSRS assignments with corresponding EHR-based definitions (any ICD-9/10 billing code).

Among the remaining 22% of participants who denied SB and ideation (N = 2,050) at baseline, fewer than 5% had a related ICD-9/10 code that predated their in-person assessment. A total of 85 participants (1%) denied SB but had a previously recorded ICD-9/10 code for attempt; and 30 (0.35%) with an EHR-documented attempt had also denied prior ideation.

Three hundred and eighty-nine (8%) participants who self-endorsed prior SB had a documented suicide attempt subsequent to their in-person C-SSRS assessment, whereas only 27 (1%) participants who denied both SB and ideation later attempted suicide based on their EHR information in the 8-12 years following their initial assessment.

### Associations between EHR-derived phenotypes and suicidal behaviors

Figure 2 displays associations between SB and 1,650 phecodes in 17 disease categories. Self-reported SB corresponded to more than six-fold increased odds of a diagnosed suicide attempt or self-inflicted injury (OR=6.56, 95% CI:4.86, 9.05; *P=*1.55×10^−32^); and 2-3-fold increases for diagnoses of mood disorders, poisonings by psychotropic agents, diagnoses of PTSD and SUDs; smaller associations were found for physical conditions including asthma, pain, sleep disorders, GERD, headache, and erectile dysfunction. Ideation at the baseline assessment (in the absence of SB) was also associated with increased odds of a diagnosed suicide attempt, personality disorders, SUDs, and PTSD, but to a lesser degree than behavior (Table S5).

**Figure 2.**
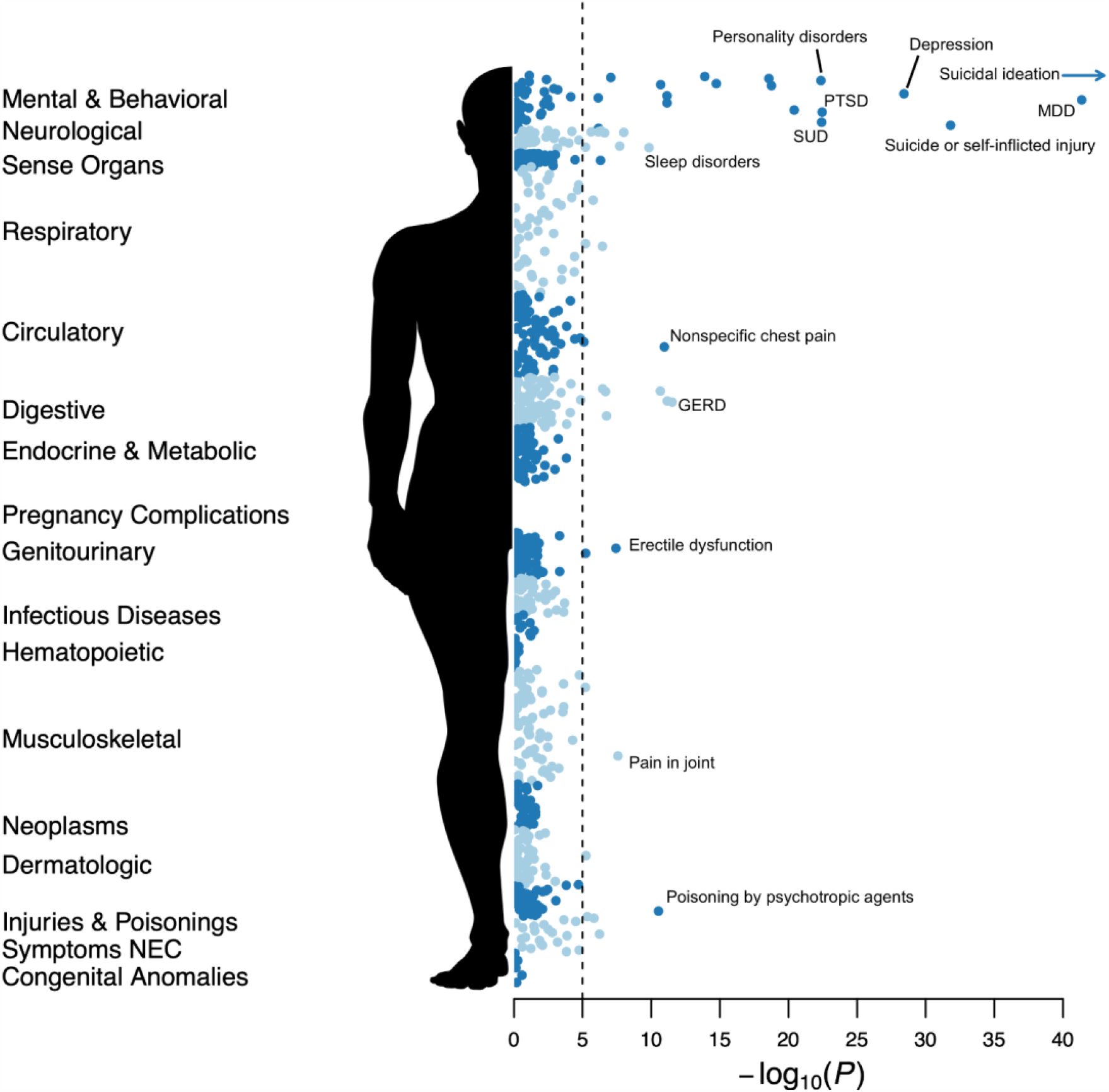
PheWAS for suicidality phenotypes in CSP #572. PheWAS results for suicidal behaviors across 1,000 disease categories. The dotted line indicates an approximate Bonferroni adjusted p-value threshold for the number of Phecodes tested.

Analogous tests using individuals’ median observed values for selected laboratory tests yielded associations between suicidal behaviors and elevated liver enzymes and lower serum potassium (Figure S1 and Table S6).

Participants reporting past SB had a higher number of unique psychiatric comorbidities compared to individuals who reported ideation only (β=0.19, 95% CI: 0.16, 0.22; *P=*5.5×10^−33^) or neither (β=0.275, 95% CI: 0.24, 0.31; *P=*3.4×10^−53^).

We also examined whether participants who self-reported SB were more likely to have been prescribed selected classes of psychotropic medications, and among those who had, whether duration of treatment was associated with increased or decreased risk of SB (Figure 3). Lifetime antidepressant treatment (any) yielded the strongest associations with SB, in particular selective serotonin reuptake inhibitors (SSRIs) (OR=3.27, 95% CI: 2.90, 3.68; *P=*1.50×10^−85^), which were robustly associated in both the SCZ and BPI subsamples (Table S7), as were so-called atypical antidepressants such as bupropion, trazodone, nefazodone, and mirtazapine (OR=3.0, 95% CI: 2.61, 3.44; *P=*1.14×10^−54^), selective norepinephrine reuptake inhibitors (SNRIs) (OR=2.54, 95% CI:2.13, 2.91; *P=*4.11×10^−40^), and tricyclic antidepressants (OR=1.81, 95% CI: 1.60, 2.07; *P=*1.30×10^−19^).

**Figure 3.**
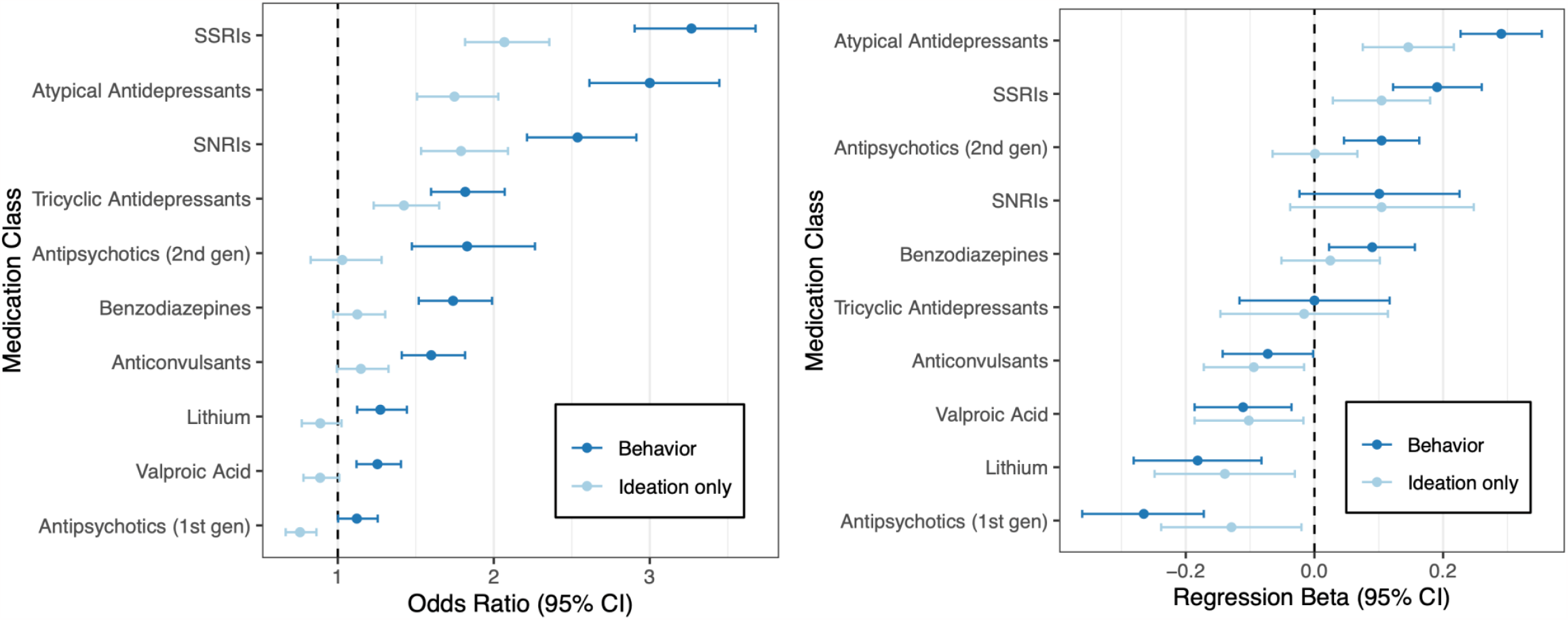
Associations of self-reported suicidality with pharmacological treatment history. (*left*) Estimated ORs for suicidal behaviors or ideation predicting lifetime treatment with selected classes of medication. (*right*) For the same medications, effect sizes for the total number of prescribed treatment-days are shown.

Self-reported SB were associated with longer duration of treatment with SSRIs (β=0.191, 95% CI: 0.1224, 0.2596; *P=*5.5×10^−8^) and atypical antidepressants (β=0.29, 95% CI:0.227, 0.353; *P=*2.09×10^−19^). Among BPI patients, SB were associated with longer treatment with second generation antipsychotics (β=0.76, 95% CI: 0.088, 0.264; *P=*9.13×10^−5^), while more days on lithium was associated with lower odds (β=-0.196, 95% CI: -0.308, -0.084; *P=*6.34×10^−4^) (Table S8).

### Associations of PGS with suicidal behavior

Figure 4 displays associations between selected PGS and SB or ideation among EA and AA participants (Tables S9 through S14). Among EA participants, SB was associated with PGS for externalizing behaviors (OR=1.203, 95% CI: 1.11, 1.31; *P*=1.23×10^−5^)(9), suicide attempt (OR=1.19, 95% CI: 1.09, 1.29; *P*=4.5×10^−5^), smoking initiation (i.e., ever/never smoked) (OR=1.19, 95% CI: 1.10, 1.30; *P*=2.77×10^−5^) (35), and major depression (OR=1.15, 95% CI: 1.06, 1.25; *P*=7.5×10^−4^) (5). The tested PGS largely did not replicate in AA participants, likely due to the fact that discovery samples were of primarily EA and PGS tend to underperform when there are discrepancies between the ancestries of the discovery and target samples (13, 48, 49). However, the externalizing behaviors PGS was nominally associated with SB (*P*<0.05) and most of the observed associations (∼75%) were in the same direction.

**Figure 4.**
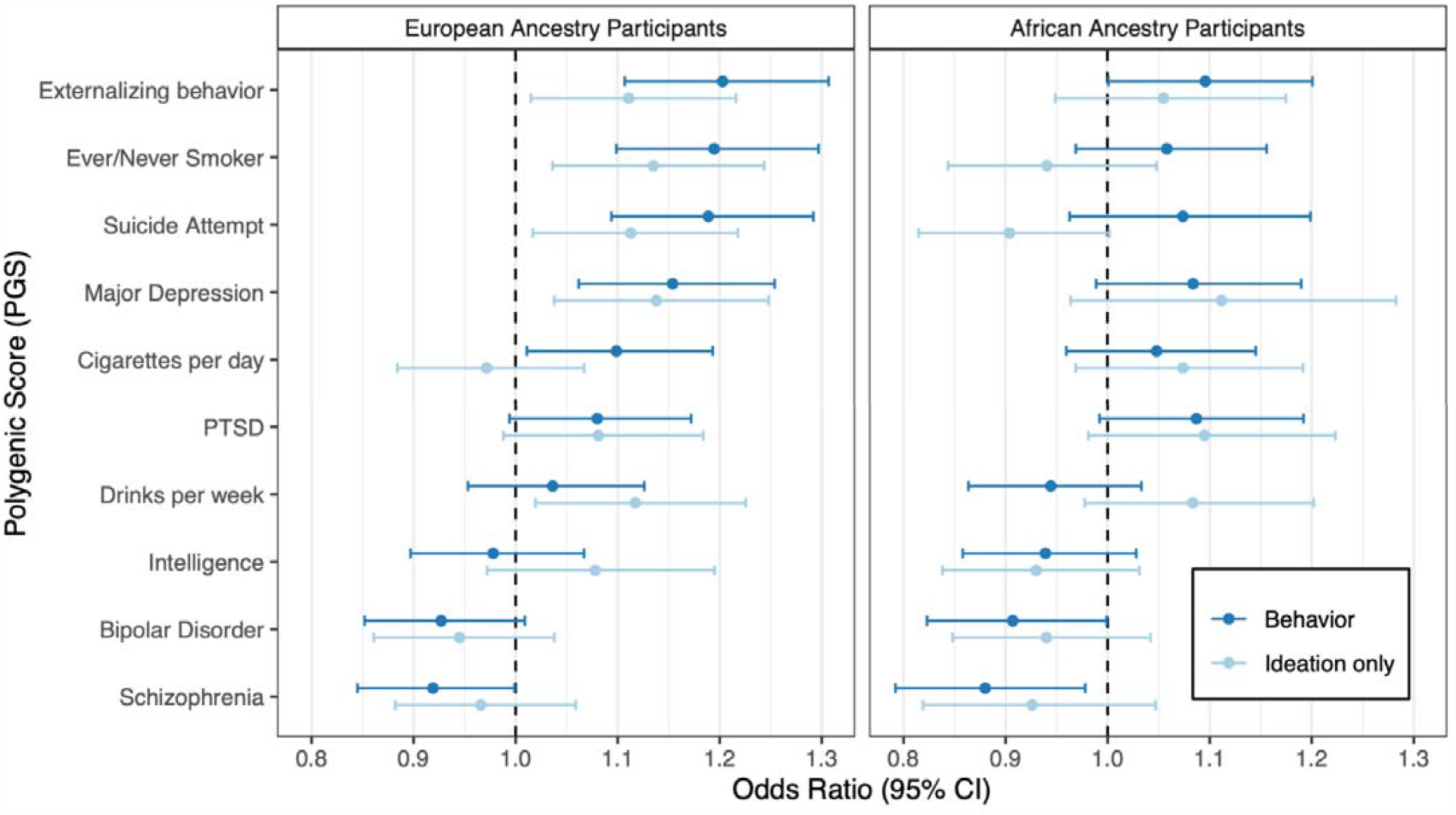
Associations of PGS with suicidality behaviors in SCZ and BPI. For each polygenic score, the odds ratio (per unit increase) corresponding to the best-performing *P*-value threshold is shown. Displayed results are based on the EA cohort, and adjusted for diagnosis, sex, and age.

**Figure 5.**
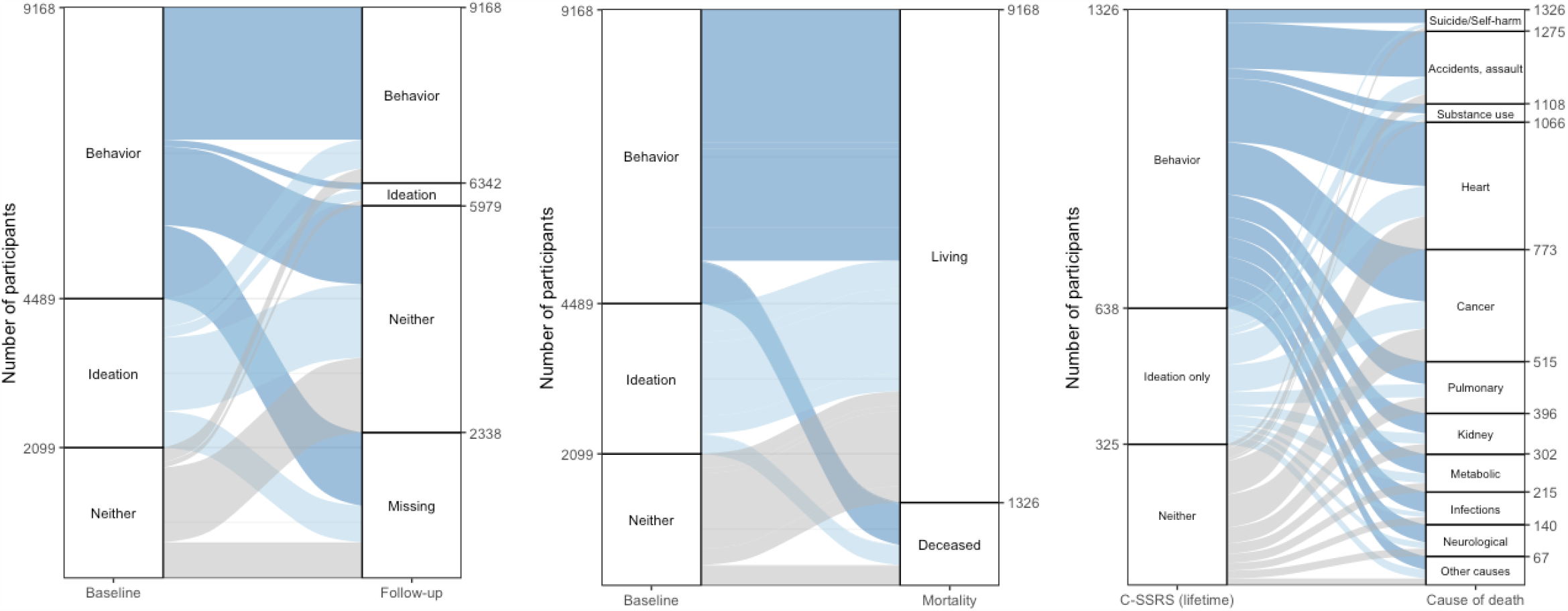
Comparisons with follow-up screenings for suicidality, mortality, and attributed cause of death. In each panel, tabular data indexing group membership and transitions are displayed as an alluvial plot. (*left*) Baseline C-SSRS from CSP #572 interviews are compared with follow-up C-SSRS screenings; “missing” indicates that additional C-SSRS data was not available in the EHR. (*middle*) for baseline C-SSRS assessments, the number of living or deceased participants is shown. (*right*) For CSP #572 participants who are deceased at the time of writing, lifetime C-SSRS-based outcomes are compared with attributed COD.

When we re-analyzed phecodes that were significantly associated with SB, incorporating the externalizing PGS as an additional predictor, we observed improved model fit and predictive value for several SUD-related traits; for tobacco-use disorder this effect was replicable in both EA and AA cohorts (Figure 4).

In this highly selected sample, we were underpowered to estimate the SNP-*h*^2^ of SB (Table S14). Only for our analysis of EA participants did the GCTA model converge, yielding a non-significant estimate of 0.063 (95% CI: -0.134, 0.261; *P*=2.73×10^−1^), albeit one that overlaps with published estimates of 4% (30).

### Exploratory Genome-wide Association Studies

No single SNP attained genome-wide significance in our primary GWAS of SB in CSP #572 (Tables S16-20; Figures S3-4). Meta-analyzing our results with published results for suicide attempts in BPI generated additional support for associations with SNPs at 4p11.21 (OR = 1.25, *P*=3.17×10^−10^) (5), (see supplementary Tables S21-24; Figure S5 for details).

A recently reported association at 7q31.2 (30) that replicated in the broader MVP sample did not replicate in CSP #572 (OR = 1.05, 95% CI: 0.95,1.16; *P*=0.285). Remarkably, the risk-increasing allele (rs62474683) had a higher frequency among CSP #572 participants who denied past suicidality than participants who endorsed past suicidality (∼58% and 52%, respectively).

Next, we imputed TWAS results directly from summary statistics of our primary GWAS and meta-analyses (Tables S25-31). In our analysis of AA participants, increased expression of *TRIM4* (7q22.1) in substantia nigra, amygdala, caudate nucleus, frontal cortex, spinal cord, and adrenal and pituitary glands was associated with SB (*q*_tissue_<0.05). The meta-analysis of CSP #572 and PGC suicide attempt GWAS summary statistics highlighted the differential expression of two genes on 17q11.2; predicted higher *CRLF3* expression in putamen and amygdala and lower *TIAF1* expression in eight CNS tissues and adrenal, pituitary, and thyroid glands survived a second FDR correction across tissues (Table S32).

### Follow-up screenings and subsequent mortality

Among 3,502 participants who self-reported SB at baseline and with available follow-up data, 2,138 (60.9%) reaffirmed this at least once during routine clinical screenings. Among 1,806 participants who reported ideation only and 1,522 who reported neither, 460 (25.5%) and 235 (15.4%) later reported SB, respectively.

Among 9,168 participants included in this analysis, 1,326 (14.5%) were deceased at the time of writing. The leading attributed causes of death were cardiovascular disease (293; 22.1%), cancer (258; 19.5%), and accidents (167, 12.6%). There were 51 (3.8%) recorded deaths by suicide. Having a SCZ diagnosis (versus BPI) was associated with increased risk of overall mortality (OR=1.31, 95% CI: 1.15, 1.50; *P*=3.55×10^−5^) while self-reported SB at baseline were not (OR=1.18, 95% CI: 0.96, 1.30; *P*=1.45×10^−1^).

However, among deceased participants, those who died by suicide or accident were more likely to have reported SB at baseline (OR=1.68, 95% CI: 1.10, 2.63; *P*=1.88×10^−2^). Notably, participants who died from suicide or self-harm were significantly younger than those with other attributed CODs (β=-0.74, 95% CI: -1.03, -0.46; *P=*3.4×10^−7^), as were those who died from accidents (β=-0.79, 95% CI: -0.98, -0.61; *P=*9.6×10^−17^).

## DISCUSSION

Building on our prior characterizations of the demographic and clinical correlates of suicidality in CSP #572 patients with SCZ and BPIr (14, 15), we have undertaken a comprehensive investigation of participants’ longitudinal EHRs including diagnosed illnesses, routine laboratory tests, prescription records, routine clinical screenings, and recorded causes of death. Additionally, we integrated newly available genomic data with these clinical data to: 1) explore associations between individual genetic variants and suicide attempt, and 2) test the predictive value of current PGS for suicide-related traits, highlighting internalizing problems (e.g., major depression) and externalizing problems (e.g., impulsivity), as possibly distinct pathways to risk.

When comparing participants’ C-SSRS responses to data from their EHRs, we found that past SB and ideation, as reported during in-person interviews, were frequently not documented in VA clinical settings. Nonetheless, self-reported SB were associated with numerous relevant psychiatric comorbidities such as PTSD, personality disorders, SUDs, and insomnia, as well as physical health problems, and accidental poisoning. Individuals who reported SB also had elevated liver enzymes, which may index medication exposures, or SUD and other poisonings.

Examining prescription records, we found that in CSP #572, patients who reported SB were more likely to have been treated with antidepressants. Among those who had received treatment with antidepressants, longer duration of treatment was positively associated with SB; in contrast, longer duration of lithium treatment was associated with a lower likelihood of self-reported SB. We did not observe that treatment with clozapine — the only medication approved by the US FDA for reducing suicide risk in SCZ (50) — was associated with lower rates of SB, but were limited by the underutilization of clozapine by practitioners; fewer than 400 SCZ patients in CSP #572 have been prescribed clozapine in VA settings.

We evaluated whether currently indexable polygenic risk for suicide-related behaviors is generalizable to this special cohort of veterans with diagnosed SMI. Patients who self-reported SB had higher PGS for externalizing, smoking, suicide attempt, and depression, extending previous findings of shared liabilities underlying suicide risk(5, 7). Patients reporting ideation-only had higher polygenic loadings than those who denied both SB and ideation, supporting ideation as a genetically informative intermediate phenotype.

Evidence in favor of specific genetic variants was more limited. We generated additional, multi-ancestry support for a 4p11.21 region downstream of *PPARGC1A* (5, 51), but not a newly identified 7q31.2 locus (30). As a secondary approach, we employed TWAS to prioritize loci based on predicted tissue-specific expression, which highlighted increased *CRLF3* and lnc-FAM72B-8, and decreased *TIAF1* and *PPP4R1*. Downregulation of *CRLF3*, which encodes cytokine receptor-like factor 3, causes abnormal neurogenesis in iPSC-derived organoid models of neurofibromatosis type 1, and is associated with more autistic features in patients carrying loss-of-function variants (52). *PP4R1*, encoding protein phosphatase 4 regulatory subunit 1, and *TIAF1*, encoding TGFB1-induced anti-apoptotic factor 1, have been implicated in recent, large-scale meta-analyses of cognitive performance (53) and major depression, respectively (54, 55).

To determine whether respondents’ accounts of past SB are stable over time and associated with an outcome of suicide, we queried electronic health records that covered a span of eight to twelve years after baseline. Although most reports of prior SB were corroborated in subsequent screenings, a large fraction of participants did not have comparable records of behaviors and ideation in their EHR. Taken together with numbers of veterans who denied any lifetime SB and ideation and later died by apparent suicides or accidents, this supports the need for direct and regular screenings by mental health professionals, especially for individuals at highest risk of suicide. For this reason, the VA and now many other hospital systems require the C-SSRS on every encounter with patients. Similarly, the absence of suicide-relevant EHR codes in the presence of repeated reports of SB on the C-SSRS suggests a need for linkage between different data sources.

### Limitations

Our primary limitation was a lack of detailed information on the dates and circumstances of participants’ prior SB. Similarly, because available EHR data is currently restricted to treatment received at VA facilities, information about behaviors and ideation occurring before or during participants’ military service is limited to self-report. We therefore cannot establish the temporality of past behaviors relative to specific ICD-9/10 codes, laboratory values, and prescription records.

A second limitation pertains to reliance on self-reported information on past suicidality. Both respondent bias — in general and in the context of SMI — and the particular settings in which routine screenings are administered may influence responses. Similarly, questions about suicidal ideation typically refer to recent weeks and months and may underestimate prevalence especially as related to episodic mood disturbances.

Finally, risk for SB is heterogeneous. This study focused on clinical characteristics of patients with SCZ or BPI. However, SB exist in patients with other psychiatric diagnoses and also in individuals who have never been diagnosed. Additionally, risk for SB is also associated with social factors (e.g., unemployment, experiences of homelessness, social support) (56). Future efforts should examine whether current results generalize to other forms of mental illness in conjunction with well-established social determinants of suicide risk.

## Conclusions

Prediction of SB is one of the most important tasks in psychiatry. This study examined whether genetic risk factors could be determined for SB regardless of diagnosis and examined other factors that determined risk. We did not detect any genome-wide significant loci. However, the sample was clinically heterogeneous in many ways. A much larger sample crossing more diagnostic boundaries may yield more positive results. Large biobanks linked to EHR have enormous potential to advance precision medicine, and synergy with more deeply characterized cohorts can enhance our characterizations of risk and resilience factors.

## Supporting information

Supplementary acknowledgements

## Data Availability

Publicly available summary statistics are available through their respective consortia websites.

## ACKNOWLEDGEMENTS

This research was supported by the Department of Veterans Affairs Cooperative Studies Program (CSP #572), the Million Veteran Program (MVP-000) and a Clinical Sciences Research and Development award (lK6BX003777). The MVP is supported by the Office of Research and Development, Department of Veterans Affairs. Full acknowledgements for CSP #572 and MVP are presented in the Supplement. The contents do not represent the views of the U.S. Department of Veterans Affairs or the United States Government.

Dr. Bigdeli was supported by a 2019 NARSAD Young Investigator Grant from the Brain & Behavior Research Foundation (#28276).

This analysis uses data from the Externalizing Consortium, supported by the National Institute on Alcohol Abuse and Alcoholism (R01AA015416), and the National Institute on Drug Abuse (R01DA050721). Additional funding for investigator effort has been provided by K02AA018755, U10AA008401, P50AA022537, and a European Research Council Consolidator Grant (647648 EdGe).

The content does not necessarily represent the views of these funding bodies. We would also like to thank the participants and employees of 23andMe, Inc. for making this work possible.

We are grateful to Drs. R. Karlsson Linnér and T.T. Mallard for sharing their script for plotting PheWAS results, which served as the basis of Figure 2.

Dr. Harvey has served as a consultant to multiple pharmaceutical companies and device manufacturers on phase 2 or 3 development; this consulting work has been determined to be unrelated to the content of the paper.

No other authors report any relevant conflicts of interest.

Larry J. Siever, original co-Principal Investigator for CSP #572, passed away in February 2021.

## REFERENCES

1. Wisqars W-BISQ: Reporting System. National Center for Injury Prevention and Control, Centers for Disease Control and Prevention (producer) Available at https://wwwcdcgov/injury/wisqars Accessed September 2020; 25

2. Thompson M, Gibbs N: More U.S. soldiers have killed themselves than have died in the Afghan war. Why can’t the Army win the war on suicide? Time 2012; 180:22–31

3. Tilstra AM, Simon DH, Masters RK: Trends in “Deaths of Despair” Among Working-Aged White and Black Americans, 1990–2017 [Internet]. American Journal of Epidemiology 2021; 190:1751– 1759Available from: http://dx.doi.org/10.1093/aje/kwab088

4. Bachmann S: Epidemiology of Suicide and the Psychiatric Perspective [Internet]. Int J Environ Res Public Health 2018; 15Available from: http://dx.doi.org/10.3390/ijerph15071425

5. Mullins N, Bigdeli TB, Børglum AD, et al.: GWAS of Suicide Attempt in Psychiatric Disorders and Association With Major Depression Polygenic Risk Scores. Am J Psychiatry 2019; 176:651–660

6. Levey DF, Polimanti R, Cheng Z, et al.: Genetic associations with suicide attempt severity and genetic overlap with major depression. Transl Psychiatry 2019; 9:22

7. Ruderfer DM, Walsh CG, Aguirre MW, et al.: Significant shared heritability underlies suicide attempt and clinically predicted probability of attempting suicide. Mol Psychiatry 2020; 25:2422– 2430

8. Docherty AR, Shabalin AA, DiBlasi E, et al.: Genome-Wide Association Study of Suicide Death and Polygenic Prediction of Clinical Antecedents. Am J Psychiatry 2020; 177:917–927

9. Karlsson Linnér R, Mallard TT, Barr PB, et al.: Multivariate analysis of 1.5 million people identifies genetic associations with traits related to self-regulation and addiction. Nat Neurosci 2021; 24:1367–1376

10. Gaziano JM, Concato J, Brophy M, et al.: Million Veteran Program: A mega-biobank to study genetic influences on health and disease. J Clin Epidemiol 2016; 70:214–223

11. Harvey PD, Siever LJ, Huang GD, et al.: The genetics of functional disability in schizophrenia and bipolar illness: methods and initial results for VA cooperative study# 572. Am J Med Genet B Neuropsychiatr Genet 2014; 165:381–389

12. Posner K, Brent D, Lucas C, et al.: Columbia-suicide severity rating scale [Internet]. New York, NY: Columbia University 2008; Available from: http://www.ccsme.org/wp-content/uploads/2017/02/Separating-the-Wheat-from-the-Chaff.pdf

13. Bigdeli T, Fanous A, Li Y, et al.: Genome-Wide Association Studies of Schizophrenia and Bipolar Disorder in a Diverse Cohort of US Veterans [Internet]. Biological Psychiatry 2020; 87:S178Available from: http://dx.doi.org/10.1016/j.biopsych.2020.02.468

14. Harvey PD, Posner K, Rajeevan N, et al.: Suicidal ideation and behavior in US veterans with schizophrenia or bipolar disorder. J Psychiatr Res 2018; 102:216–222

15. Aslan M, Radhakrishnan K, Rajeevan N, et al.: Suicidal ideation, behavior, and mortality in male and female US veterans with severe mental illness [Internet]. Journal of Affective Disorders 2020; 267:144–152Available from: http://dx.doi.org/10.1016/j.jad.2020.02.022

16. First MB, Spitzer RL, Gibbon M, et al.: User’s guide for the Structured clinical interview for DSM-IV axis I disorders SCID-I: clinician version. American Psychiatric Pub, 1997

17. Posner K, Brent D, Lucas C, et al.: Columbia-suicide severity rating scale (C-SSRS) [Internet]. New York, NY: Columbia University Medical Center 2008; 10Available from: https://vtspc.org/wp-content/uploads/2016/12/C-SSRS-LifetimeRecent-Clinical.pdf

18. Sheehan DV, Lecrubier Y, Sheehan KH, et al.: The Mini-International Neuropsychiatric Interview (M.I.N.I.): the development and validation of a structured diagnostic psychiatric interview for DSM-IV and ICD-10. J Clin Psychiatry 1998; 59 Suppl 20:22–33;quiz 34–57

19. Liao KP, Sun J, Cai TA, et al.: High-throughput multimodal automated phenotyping (MAP) with application to PheWAS. J Am Med Inform Assoc 2019; 26:1255–1262

20. Denny JC, Ritchie MD, Basford MA, et al.: PheWAS: demonstrating the feasibility of a phenome-wide scan to discover gene–disease associations. Bioinformatics 2010; 26:1205–1210[cited 2021 Feb 7]

21. Bigdeli TB, Voloudakis G, Barr PB, et al.: Penetrance and Pleiotropy of Polygenic Risk Scores for Schizophrenia, Bipolar Disorder, and Depression Among Adults in the US Veterans Affairs Health Care System [Internet]. JAMA Psychiatry 2022; Available from: https://jamanetwork.com/journals/jamapsychiatry/article-abstract/2796413

22. Klarin D, Damrauer SM, Cho K, et al.: Genetics of blood lipids among ∼300,000 multi-ethnic participants of the Million Veteran Program. Nat Genet 2018; 50:1514–1523

23. 1000 Genomes Project Consortium, Auton A, Brooks LD, et al.: A global reference for human genetic variation. Nature 2015; 526:68–74

24. Das S, Forer L, Schönherr S, et al.: Next-generation genotype imputation service and methods. Nat Genet 2016; 48:1284–1287

25. Manichaikul A, Mychaleckyj JC, Rich SS, et al.: Robust relationship inference in genome-wide association studies. Bioinformatics 2010; 26:2867–2873

26. Fang H, Hui Q, Lynch J, et al.: Harmonizing Genetic Ancestry and Self-identified Race/Ethnicity in Genome-wide Association Studies. Am J Hum Genet 2019; 105:763–772

27. Bigdeli TB, Genovese G, Georgakopoulos P, et al.: Contributions of common genetic variants to risk of schizophrenia among individuals of African and Latino ancestry [Internet]. Mol Psychiatry 2019; Available from: http://dx.doi.org/10.1038/s41380-019-0517-y

28. Peterson RE, Kuchenbaecker K, Walters RK, et al.: Genome-wide Association Studies in Ancestrally Diverse Populations: Opportunities, Methods, Pitfalls, and Recommendations. Cell 2019; 179:589–603

29. Abraham G, Inouye M: Fast principal component analysis of large-scale genome-wide data. PLoS One 2014; 9:e93766

30. Mullins N, Kang J, Campos AI, et al.: Dissecting the shared genetic architecture of suicide attempt, psychiatric disorders and known risk factors [Internet]. Biol Psychiatry 2021; Available from: https://www.sciencedirect.com/science/article/pii/S0006322321015705

31. Trubetskoy V, Pardiñas AF, Qi T, et al.: Mapping genomic loci implicates genes and synaptic biology in schizophrenia. Nature 2022; 604:502–508

32. Nievergelt CM, Maihofer AX, Klengel T, et al.: International meta-analysis of PTSD genome-wide association studies identifies sex- and ancestry-specific genetic risk loci. Nat Commun 2019; 10:4558

33. Mullins N, Forstner AJ, O’Connell KS, et al.: Genome-wide association study of more than 40,000 bipolar disorder cases provides new insights into the underlying biology. Nat Genet 2021; 53:817– 829

34. Howard DM, Adams MJ, Clarke T-K, et al.: Genome-wide meta-analysis of depression identifies 102 independent variants and highlights the importance of the prefrontal brain regions. Nat Neurosci 2019; 22:343–352

35. Liu M, Jiang Y, Wedow R, et al.: Association studies of up to 1.2 million individuals yield new insights into the genetic etiology of tobacco and alcohol use. Nat Genet 2019; 51:237–244

36. Savage JE, Jansen PR, Stringer S, et al.: Genome-wide association meta-analysis in 269,867 individuals identifies new genetic and functional links to intelligence. Nat Genet 2018; 50:912–919

37. Lee SH, Wray NR, Goddard ME, et al.: Estimating missing heritability for disease from genome-wide association studies. Am J Hum Genet 2011; 88:294–305

38. Yang J, Lee SH, Goddard ME, et al.: GCTA: a tool for genome-wide complex trait analysis. Am J Hum Genet 2011; 88:76–82

39. Purcell S, Neale B, Todd-Brown K, et al.: PLINK: a tool set for whole-genome association and population-based linkage analyses. Am J Hum Genet 2007; 81:559–575

40. Chang CC, Chow CC, Tellier LC, et al.: Second-generation PLINK: rising to the challenge of larger and richer datasets. Gigascience 2015; 4:7

41. Willer CJ, Li Y, Abecasis GR: METAL: fast and efficient meta-analysis of genomewide association scans. Bioinformatics 2010; 26:2190–2191

42. Schizophrenia Working Group of the Psychiatric Genomics, Consortium: Biological insights from 108 schizophrenia-associated genetic loci. Nature 2014; 511:421–427

43. Watanabe K, Taskesen E, van Bochoven A, et al.: Functional mapping and annotation of genetic associations with FUMA. Nat Commun 2017; 8:1826

44. Pruim RJ, Welch RP, Sanna S, et al.: LocusZoom: regional visualization of genome-wide association scan results. Bioinformatics 2010; 26:2336–2337

45. Lee CH, Eskin E, Han B: Increasing the power of meta-analysis of genome-wide association studies to detect heterogeneous effects. Bioinformatics 2017; 33:i379–i388

46. Gamazon ER, Wheeler HE, Shah KP, et al.: A gene-based association method for mapping traits using reference transcriptome data. Nat Genet 2015; 47:1091–1098

47. Barbeira AN, Dickinson SP, Bonazzola R, et al.: Exploring the phenotypic consequences of tissue specific gene expression variation inferred from GWAS summary statistics. Nat Commun 2018; 9:1825

48. Martin AR, Gignoux CR, Walters RK, et al.: Human Demographic History Impacts Genetic Risk Prediction across Diverse Populations. Am J Hum Genet 2017; 100:635–649

49. Duncan L, Shen H, Gelaye B, et al.: Analysis of polygenic risk score usage and performance in diverse human populations [Internet]. Nature Communications 2019; 10Available from: http://dx.doi.org/10.1038/s41467-019-11112-0

50. DailyMed-CLOZAPINE tablet [Internet][cited 2022 Oct 28] Available from: https://dailymed.nlm.nih.gov/dailymed/drugInfo.cfm?setid=883b5d43-0339-7dc1-f775-93791fb9b978

51. Pardiñas AF, Holmans P, Pocklington AJ, et al.: Common schizophrenia alleles are enriched in mutation-intolerant genes and in regions under strong background selection. Nat Genet 2018; 50:381–389

52. Wegscheid ML, Anastasaki C, Hartigan KA, et al.: Patient-derived iPSC-cerebral organoid modeling of the 17q11.2 microdeletion syndrome establishes CRLF3 as a critical regulator of neurogenesis. Cell Rep 2021; 36:109315

53. Lee JJ, Wedow R, Okbay A, et al.: Gene discovery and polygenic prediction from a genome-wide association study of educational attainment in 1.1 million individuals. Nat Genet 2018; 50:1112– 1121

54. Wray NR, Ripke S, Mattheisen M, et al.: Genome-wide association analyses identify 44 risk variants and refine the genetic architecture of major depression. Nat Genet 2018; 50:668–681

55. Beurel E, Toups M, Nemeroff CB: The Bidirectional Relationship of Depression and Inflammation: Double Trouble. Neuron 2020; 107:234–256

56. Blosnich JR, Montgomery AE, Dichter ME, et al.: Social Determinants and Military Veterans’ Suicide Ideation and Attempt: a Cross-sectional Analysis of Electronic Health Record Data. J Gen Intern Med 2020; 35:1759–1767

