## Supplementary acknowledgements for "Correlates of suicidal behaviors and genetic risk among United States veterans with schizophrenia or bipolar I disorder"

#### The CSP #572 study team

**Planning Committee:** M. Aslan, M. Antonelli, M. de Asis, M. S. Bauer, M. Brophy, J. Concato, F. Cunningham, R. Freedman, M. Gaziano, T. Gleason, P. D. Harvey, G. Huang, J. Kelsoe, T. Kosten, T. Lehner, J. B. Lohr, S. R. Marder, P. Miller, T.J. O’Leary, T. Patterson, P. Peduzzi, R. Przygodzki, L. Siever, P. Sklar, S. Strakowski, H. Zhao.

**Executive Committee:** M. Brophy, J. Concato, A.H. Fanous, M. Gaziano, P.D. Harvey, T. Kosten, A. Malhotra, S. Mane, T. Bigdeli, P. Sklar, L. Siever, H. Zhao.

**Study Chairs’ Offices:** VA Healthcare System, Bronx, NY: L. Siever (Co-Chair), M. Corsey, L. Zaluda. VA Healthcare System, Miami, FL: P.D. Harvey (Co-Chair), J. Johnson.

**CSP Epidemiology Centers:** VA Clinical Epidemiology Research Center CERC, VA Connecticut Healthcare System, West Haven, CT, included J. Concato Director, Methodological Co-Principal Proponent, M. Aslan, D. Cavaliere, V. Jeanpaul, A. Maffucci, L. Mancini; the Massachusetts Veterans Epidemiology Research and Information Center MAVERIC, VA Boston Healthcare System, Jamaica Plain, MA, included M. Gaziano Director, Methodological Co-Principal Proponent, J. Deen, G. Muldoon, S. Whitbourne.

**Study Sites:** *Albuquerque*: J**. Canive,** L. Adamson, L. Calais, G. Fuldauer, R. Kushner, G. Toney, M. Lackey, A. Mank, N. Mahdavi, G. Villarreal. *Atlanta*: **E. C. Muly,** F. Amin, M. Dent, J. Wold. *Baltimore*: **B. Fischer,** A. Elliott, C. Felix, G. Gill. *Birmingham*: **P. E. Parker**, C. Logan, J. McAlpine. Boston/*Brockton*: **L.E. DeLisi,.** *Charleston*: **M.B. Hammer,** D. Agbor-Tabie, W. Goodson. *Cincinnati*: **M. Aslam,** M. Grainger, Neil Richtand, Alexander Rybalsky. *Houston*: **R. Al Jurdi**, E. Boeckman, T. Natividad, D. Smith, M. Stewart, S. Torres, Z. Zhao. *Indianapolis*: **A. Mayeda,** A. Green, J. Hofstetter, S. Ngombu, M. K. Scott, A. Strasburger, J. Sumner. *Little Rock*: **G. Paschall**, J. Mucciarelli, R. Owen, S. Theus, D. Tompkins. *Long Beach*: **S.G. Potkin,** C. Reist, M. Novin, S. Khalaghizadeh. *Miami*: **R. Douyon**, J. Johnson, N. Kumar, B. Martinez. *Minneapolis*: **S.R. Sponheim,** T.L. Bender, H.L. Lucas, A.M. Lyon, M.P. Marggraf, L.H. Sorensen, C.R. Surerus. *Montrose***: C. Sison,** J. Amato, D.R. Johnson, N. Pagan-Howard. *New York Harbor*: **L.A. Adler,** S. Alperin, T. Leon. *Northampton*: **K.M. Mattocks**, N. Araeva, J.C. Sullivan. *Palo Alto*: **T. Suppes,** K. Bratcher, L. Drag, E.G. Fischer, L. Fujitani, S. Gill, D. Grimm, J. Hoblyn, T. Nguyen, E. Nikolaev, L. Shere, R. Relova, A. Vicencio, M. Yip. *Philadelphia***: I. Hurford,** S. Acheampong, G. Carfagno. *Pittsburgh*: **G.L. Haas**, C. Appelt, E. Brown, B. Chakraborty, E. Kelly, G. Klima, S. Steinhauer. *Salisbury*: **R.A. Hurley,** R. Belle, D. Eknoyan, K. Johnson, J. Lamotte. *San Diego*: **E. Granholm**, K. Bradshaw, J. Holden, R. H. Jones, T. Le, I.G. Molina, M. Peyton, I. Ruiz, L. Sally. *Tacoma*: **A. Tapp,** S. Devroy, V. Jain, N. Kilzieh, L. Maus, K. Miller, H. Pope, A. Wood. *Temple*: **E. Meyer,** P. Givens, P. B. Hicks, S. Justice, K. McNair, J.L. Pena, D.F. Tharp. *Tuscaloosa*: **L. Davis,** M. Ban, L. Cheatum, P. Darr, W. Grayson, J. Munford, D. Smith, B. Whitfield, E. Wilson. *Washington DC*: **A.H. Fanous,** S.E. Melnikoff, B.L Schwartz, M.A. Tureson. *West Haven*: D. D’Souza, K. Forselius, M. Ranganathan, L. Rispoli.

**Albuquerque, NM, CSP Coordinating Center**: M. Sather Director, C. Colling, C. Haakenson, D. Krueger.

**VA Office of Research and Development**: T. O’Leary Chief Research and Development Officer, G. Huang Director, Cooperative Studies Program, T. Gleason Director, Clinical Science Research and Development Service, R. Przygodzki Associate Director for Genomic Medicine, and Acting Director of Biomedical Laboratory Research and Development Service, S. Muralidhar Senior Scientific Program Manager Genomic Medicine Program, Biomedical and Clinical R&D Services.

#### Million Veteran Program: Consortium Acknowledgement for Manuscripts

**MVP Executive Committee**

- Co-Chair: J. Michael Gaziano, M.D., M.P.H.

- Co-Chair: Rachel Ramoni, D.M.D., Sc.D.

- Jim Breeling, M.D. ex-officio

- Kyong-Mi Chang, M.D.

- Grant Huang, Ph.D.

- Sumitra Muralidhar, Ph.D.

- Christopher J. O’Donnell, M.D., M.P.H.

- Philip S. Tsao, Ph.D.

**MVP Program Office**

- Sumitra Muralidhar, Ph.D.

- Jennifer Moser, Ph.D.

**MVP Recruitment/Enrollment**

- Recruitment/Enrollment Director/Deputy Director, Boston

- Stacey B. Whitbourne, Ph.D.; Jessica V. Brewer, M.P.H.

- MVP Coordinating Centers

o Clinical Epidemiology Research Center CERC, West Haven – John Concato, M.D., M.P.H.

o Cooperative Studies Program Clinical Research Pharmacy Coordinating Center, Albuquerque - Stuart Warren, J.D., Pharm D.; Dean P. Argyres, M.S.

o Genomics Coordinating Center, Palo Alto – Philip S. Tsao, Ph.D.

o Massachusetts Veterans Epidemiology Research Information Center MAVERIC, Boston - J. Michael Gaziano, M.D., M.P.H.

o MVP Information Center, Canandaigua – Brady Stephens, M.S.

- Core Biorepository, Boston – Mary T. Brophy M.D., M.P.H.; Donald E. Humphries, Ph.D.

- MVP Informatics, Boston – Nhan Do, M.D.; Shahpoor Shayan

- Data Operations/Analytics, Boston – Xuan-Mai T. Nguyen, Ph.D.

**MVP Science**

- Genomics - Christopher J. O’Donnell, M.D., M.P.H.; Saiju Pyarajan Ph.D.; Philip S. Tsao, Ph.D.

- Phenomics - Kelly Cho, M.P.H, Ph.D.

- Data and Computational Sciences – Saiju Pyarajan, Ph.D.

- Statistical Genetics – Elizabeth Hauser, Ph.D.; Yan Sun, Ph.D.; Hongyu Zhao, Ph.D.

**MVP Local Site Investigators**

- Atlanta VA Medical Center Peter Wilson - Bay Pines VA Healthcare System Rachel McArdle

- Birmingham VA Medical Center Louis Dellitalia

- Cincinnati VA Medical Center John Harley

- Clement J. Zablocki VA Medical Center Jeffrey Whittle

- Durham VA Medical Center Jean Beckham

- Edith Nourse Rogers Memorial Veterans Hospital John Wells

- Edward Hines, Jr. VA Medical Center Salvador Gutierrez

- Fayetteville VA Medical Center Gretchen Gibson

- VA Health Care Upstate New York Laurence Kaminsky

- New Mexico VA Health Care System Gerardo Villareal

- VA Boston Healthcare System Scott Kinlay

- VA Western New York Healthcare System Junzhe Xu

- Ralph H. Johnson VA Medical Center Mark Hamner

- Wm. Jennings Bryan Dorn VA Medical Center Kathlyn Sue Haddock

- VA North Texas Health Care System Sujata Bhushan

- Hampton VA Medical Center Pran Iruvanti

- Hunter Holmes McGuire VA Medical Center Michael Godschalk

- Iowa City VA Health Care System Zuhair Ballas

- Jack C. Montgomery VA Medical Center Malcolm Buford

- James A. Haley Veterans’ Hospital Stephen Mastorides

- Louisville VA Medical Center Jon Klein

- Manchester VA Medical Center Nora Ratcliffe

- Miami VA Health Care System Hermes Florez

- Michael E. DeBakey VA Medical Center Alan Swann

- Minneapolis VA Health Care System Maureen Murdoch

- N. FL/S. GA Veterans Health System Peruvemba Sriram

- Northport VA Medical Center Shing Shing Yeh

- Overton Brooks VA Medical Center Ronald Washburn

- Philadelphia VA Medical Center Darshana Jhala

- Phoenix VA Health Care System Samuel Aguayo

- Portland VA Medical Center David Cohen

- Providence VA Medical Center Satish Sharma

- Richard Roudebush VA Medical Center John Callaghan

- Salem VA Medical Center Kris Ann Oursler

- San Francisco VA Health Care System Mary Whooley

- South Texas Veterans Health Care System Sunil Ahuja

- Southeast Louisiana Veterans Health Care System Amparo Gutierrez

- Southern Arizona VA Health Care System Ronald Schifman

- Sioux Falls VA Health Care System Jennifer Greco

- St. Louis VA Health Care System Michael Rauchman

- Syracuse VA Medical Center Richard Servatius

- VA Eastern Kansas Health Care System Mary Oehlert

- VA Greater Los Angeles Health Care System Agnes Wallbom

- VA Loma Linda Healthcare System Ronald Fernando

- VA Long Beach Healthcare System Timothy Morgan

- VA Maine Healthcare System Todd Stapley

- VA New York Harbor Healthcare System Scott Sherman

- VA Pacific Islands Health Care System Gwenevere Anderson

- VA Palo Alto Health Care System Philip Tsao

- VA Pittsburgh Health Care System Elif Sonel

- VA Puget Sound Health Care System Edward Boyko

- VA Salt Lake City Health Care System Laurence Meyer

- VA San Diego Healthcare System Samir Gupta

- VA Southern Nevada Healthcare System Joseph Fayad

- VA Tennessee Valley Healthcare System Adriana Hung

- Washington DC VA Medical Center Jack Lichy

- W.G. Bill Hefner VA Medical Center Robin Hurley

- White River Junction VA Medical Center Brooks Robey

- William S. Middleton Memorial Veterans Hospital Robert Striker

### Supplementary Figures

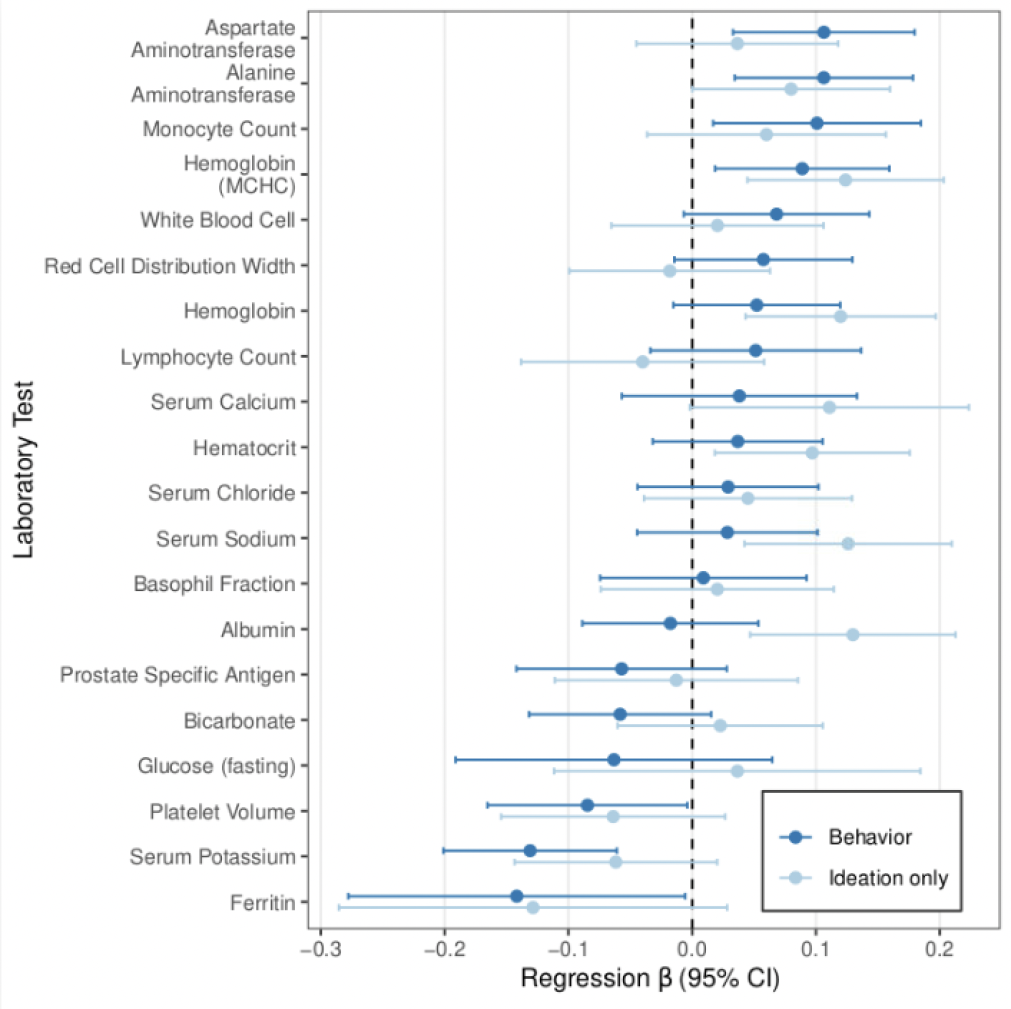

#### **Figure S1. LabWAS for suicidality phenotypes in CSP #572.**

Estimated effects sizes on common laboratory values. Displayed labs were nominally significant (P<0.05) in combined or within-disorder tests.

#### **Figure S2. Regional association of MIR548AJ2 with SB in BPI cases.**

*(top)* Regional association plot displaying strength of LD with the lead variant at *MIR548AJ2*. (*bottom*) Forest plot of effect sizes in CSP #572, PGC, and ISGC GWAS. *AF_case* and *AF_cont* are the tested allele frequencies (G) in cases and controls; *INFO* is the statistical imputation information; *OR* and *SE* are the estimated allelic odds ratio and its standard error; and *Cases* and *Controls* are the corresponding sample sizes.

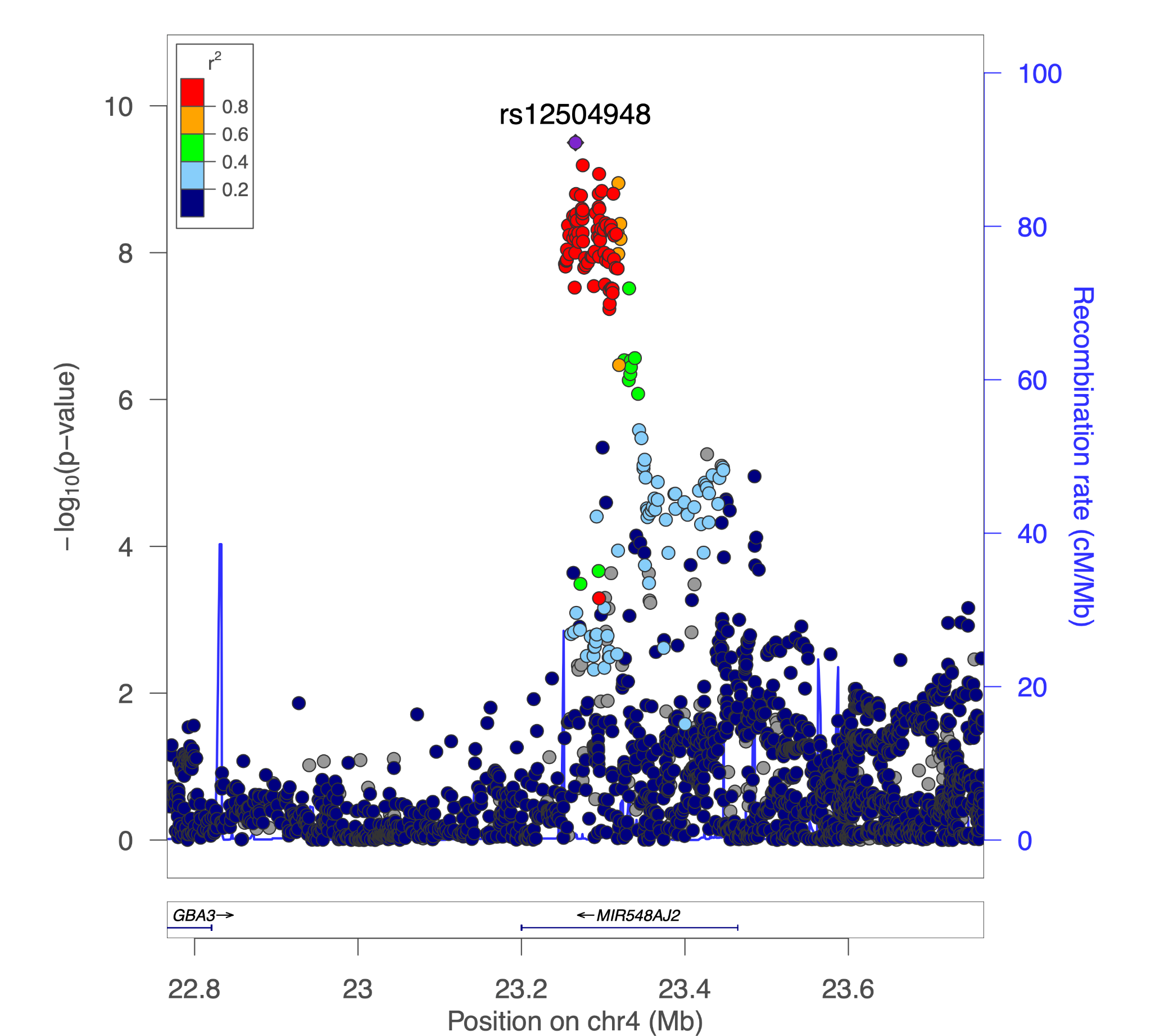

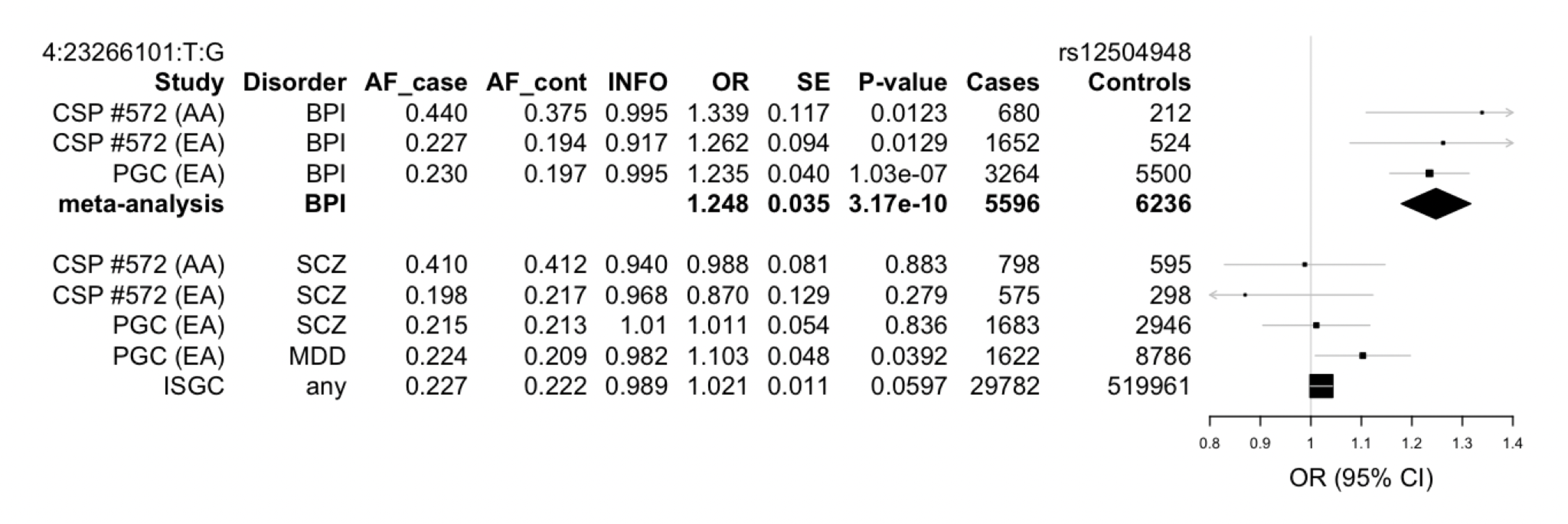
